# Early initiation of polymyxin B hemoperfusion therapy for cancer patients with refractory septic shock

**DOI:** 10.1101/2023.09.21.23295886

**Authors:** Jae Hoon Lee, Won Ho Han

**Affiliations:** Critical Care Medicine, National Cancer Center, 323 Ilsan-ro, Ilsandong-gu, Goyang-si 10408, Gyeonggi-do, Republic of Korea

**Keywords:** sepsis, 28-day mortality, risk factors, hemodynamics, respiratory function, organ failure

## Abstract

**Purpose:** In this study, we analyzed correlations between 28-day mortality and hemodynamic changes, measured using polymyxin B-immobilized fiber column direct hemoperfusion initiation time, in patients with cancer with refractory septic shock.

**Materials and methods:** We retrospectively analyzed 45 patients with cancer who received polymyxin B-immobilized fiber column direct hemoperfusion due to refractory septic shock. Patients were categorized into early (<12 h between refractory septic shock and initiation of polymyxin B-immobilized fiber column direct hemoperfusion) and late (>12 h) initiation groups. Changes in vasoactive inotrope scores, sequential organ failure assessment scores, and PaO_2_/FiO_2_ ratios before and 24 h after polymyxin B-immobilized fiber column direct hemoperfusion, were compared.

**Results:** Univariable analysis showed that 28-day mortality risk was associated with diabetes mellitus (odds ratio=3.081; 95% confidence interval =1.290–7.360; *p*=0.011), lactic acid (odds ratio=1.010; 95% confidence interval =1.005– 1.014; *p*<0.0001), and sequential organ failure assessment score (odds ratio=1.190; 95% confidence interval =1.044–1.357; *p*=0.009). Multivariable analysis showed that 28-day mortality risk was associated with diabetes mellitus (odds ratio=2.718; 95% confidence interval =1.013–7.291; *p*=0.047), early initiation (odds ratio=0.268; 95% confidence interval =0.094–0.765; *p*=0.014), and lactic acid (odds ratio=1.009; 95% confidence interval =1.004– 1.014; *p*<0.0001). Overall survival was slightly higher in the early than in the late initiation group (*p*=0.0515). Comparisons of variables before and 24 h after polymyxin B-immobilized fiber column direct hemoperfusion revealed that vasoactive inotrope scores decreased in both the early and late groups (Δ318 *vs.* Δ114; *p*=0.001 and *p*=0.005, respectively), whereas the PaO_2_/FiO_2_ ratio slightly increased (Δ127.5 *vs.* Δ95.6; *p*=0.350 and *p*=0.390, respectively) over time.

**Conclusions:** In patients with cancer with refractory septic shock, early initiation of polymyxin B-immobilized fiber column direct hemoperfusion reduced inotrope-vasopressor requirement and 28-day mortality.

## Introduction

Sepsis causes life-threatening organ dysfunction through dysregulation of host responses to infection [1]. Eleven million of the 49 million people that contract sepsis globally each year die, equivalent to a total global mortality of 19.7%. However, sepsis-related mortality has shown a decreasing trend with an increase in early detection and improvements in therapeutic strategies [2]. Sepsis is a significant risk factor in immunocompromised patients with cancer undergoing treatment, as well as in those with a known poor prognosis. The incidence of sepsis is 3- to 5-fold higher in patients with cancer, at 16.4 cases per 1,000 persons, than in those without. Furthermore, the proportion of sepsis-related mortality is approximately 3-fold higher than that of annual cancer deaths (10% *vs.* 37.8%), although mortality rates vary across cancer types [3].

Polymyxin B-immobilized fiber column direct hemoperfusion (PMX-DHP) is a potential therapy developed in 1994 to reduce blood endotoxin levels and treat septic shock, which effectively enhances hemodynamics and respiratory functions in patients with septic shock, caused by intra-abdominal infection [4]. However, whether it contributes to survival and prevents organ failure is controversial, and there are no established guidelines on the best time to perform PMX-DHP [5-8].

To date, studies on PMX-DHP have mainly targeted patients with no cancer, and there is a scarcity of literature on patients with cancer. This prompted the present investigation, in which we analyzed the correlation between PMX-DHP initiation time and 28-day mortality in patients with cancer with refractory septic shock. Additionally, we aimed to verify whether hemodynamics, sequential organ failure assessment (SOFA) scores, and oxygenation levels improve following PMX-DHP treatment.

## Materials and methods

This single-center, retrospective study was conducted in 45 patients with cancer aged ≥18 years, who received PMX-DHP to treat septic shock, at the intensive care unit of our institution between October 2019 and March 2023. Septic shock was identified based on the criteria established for identification of sepsis and septic shock (Sepsis-3), including the requirement of vasopressors to maintain a mean arterial pressure (MAP) of ≥65 mm Hg and a serum lactate level of >2 mmol/L in the absence of hypovolemia according [1]. PMX-DHP was administered to patients who voluntarily consented to the treatment. Refractory septic shock was defined as the requirement of ≥0.5 µg/kg/min of norepinephrine or an additional dose of vasopressin to maintain MAP of >65 mmHg [9]. We administered IV hydrocortisone at a dose of 200 mg/d, provided as 50 mg intravenously every 6 hours, to all patients diagnosed with refractory septic shock.

For PMX-DHP, the blood flow rate was set at 100 mL/min and controlled by the attending physician depending on the patient’s state to maintain a steady rate. For anticoagulation, 20–40 mg/h of nafamostat mesylate, a synthetic serine protease inhibitor, was administered. The duration of PMX-DHP was 2 h or more; the infusion time and frequency were determined by the attending physicians [10,11]. Patients who received PMX-DHP treatment within 12 h from the diagnosis of refractory septic shock were assigned to the “early initiation” group, while those who were treated after 12 h or more had elapsed since their diagnosis were assigned to the “late initiation” group.

The following patient characteristics were entered as variables for the analyses: age, sex, underlying disease (diabetes mellitus (DM), hypertension, cerebrovascular accident, chronic kidney disease, and chronic obstructive pulmonary disease), cancer type (intra-abdominal/genitourinary/lung/others), pre-PMX-DHP laboratory data (absolute neutrophil count, platelet count, C-reactive protein, lactic acid levels, procalcitonin, creatinine levels, PaO_2_/FiO_2_ ratio), severity (acute physiology and chronic health evaluation III (APACHE III)), SOFA score, PMX-DHP frequency, vasoactive inotrope scores (VIS), primary infection site (lung, abdominal, genitourinary, others), microbiological culture, mechanical ventilator, intervention, surgical intervention, and 28-day mortality.

VIS were measured as follows [12]:

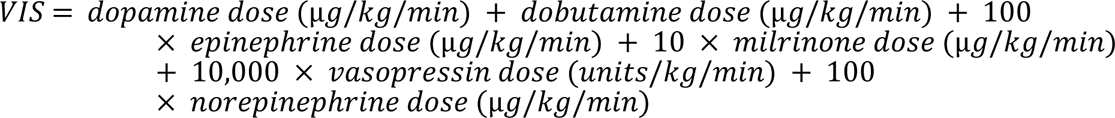

Using patient data, correlations of the aforementioned variables with PMX-DHP initiation time (early initiation group <12 h; late initiation group >12 h) were assessed by calculating differences in values reported before and 24 h after PMX-DHP. This study was approved by the Institutional Review Board of the National Cancer Center (Approval No.: NCC2023-0170). The data access period for the manuscript is from July 3, 2023, the date IRB approval, to July 21, 2023. The requirement for the acquisition of informed consent from patients was waived owing to the retrospective nature of this study.

### Statistical analysis

Continuous variables are expressed as mean ± standard deviation or median (minimum-maximum or range). Categorical variables were compared between groups using the Pearson χ^2^ and Fisher’s exact test, and continuous variables were compared using *t*-tests for parametric and the Wilcoxon rank sum test for nonparametric data. Statistical significance was set at *p*<0.05. The final set of variables in the multivariable model were selected via backward elimination in the univariable analysis, with *p* <0.1. All statistical analyses were performed using SAS® version 9.4 for Windows® (SAS Institute, Cary, NC, USA).

## Results

### Characteristics of the patient group undergoing PMX-DHP treatment

A total of 45 patients received PMX-DHP for refractory septic shock. Patients’ mean age was 64.4 years, and 24 patients (53.5%) died within 28 days due to multi-organ failure caused by septic shock. The most frequent type of cancers were genitourinary (53.3%), intra-abdominal cancers (31.1%), and lung cancer (8.9%). The most frequent primary infection site was the abdomen (64.4%), followed by the lung (20.0%), and the genitourinary tract (11.1%). Gram-negative bacteria were the most common microorganism (62.5%). The median inotropes-vasopressor requirement (VIS) to maintain an MAP ≥65 mmHg was 289.7 (9–913). The mean APACHE III score before PMX-DHP was 93.9±29.2, while the mean SOFA score was 13.2±3.3. Additionally, 41 patients underwent mechanical ventilation (91.1%) and the median PaO_2_/FiO_2_ ratio was 141.2 (20.2–534.3) (Table 1).

**Table 1.**
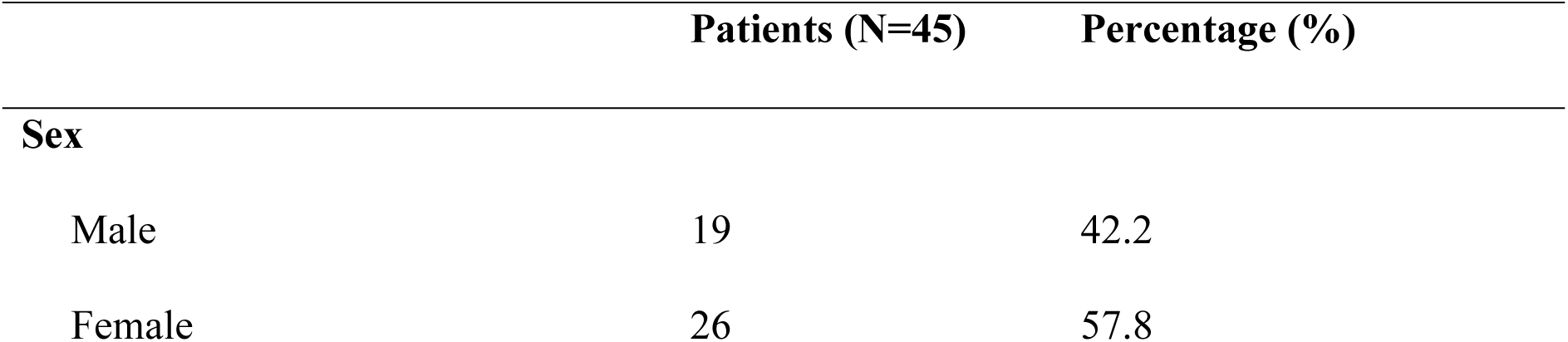
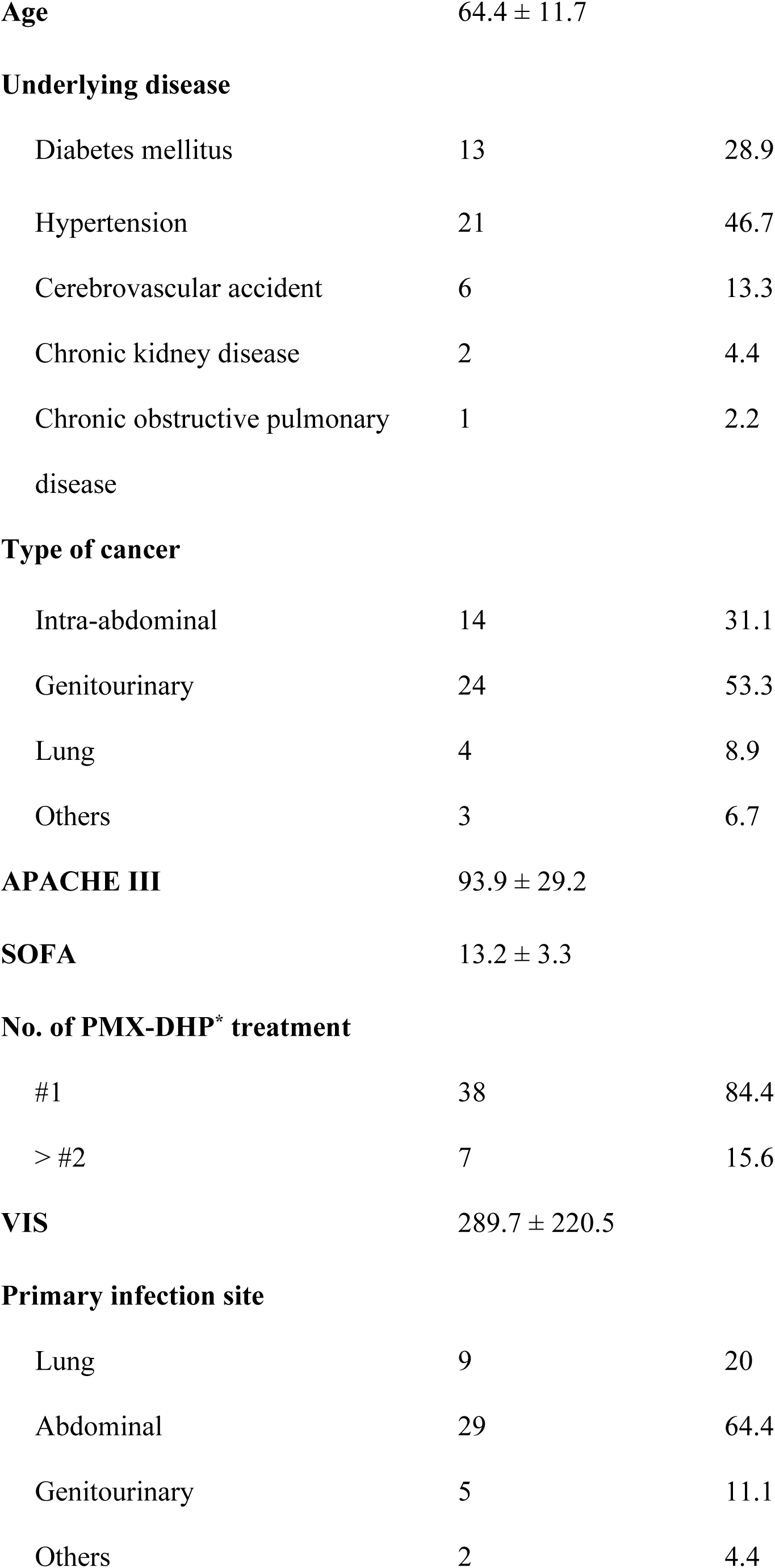
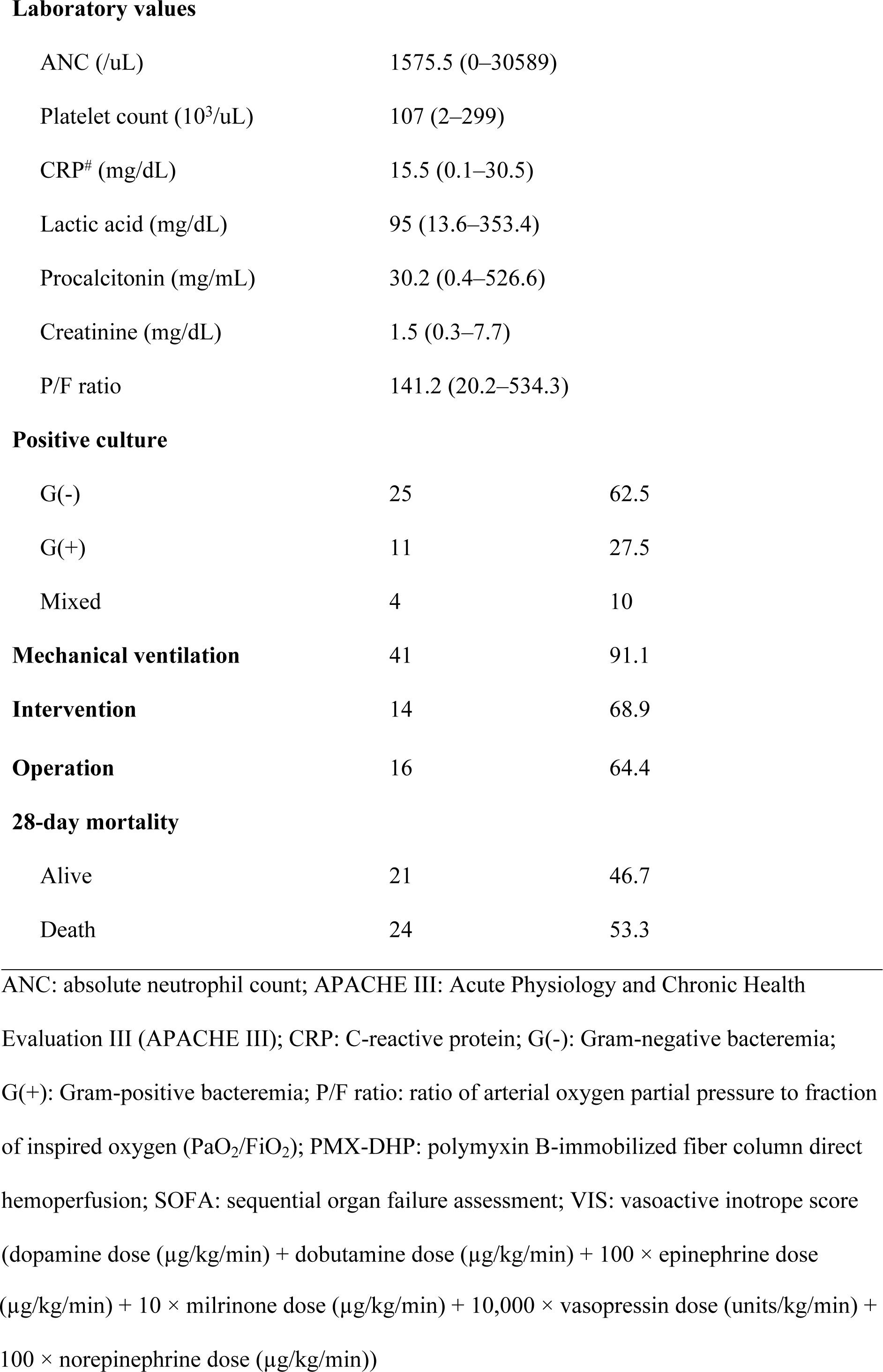
Characteristics of patients treated with PMX-DHP.

|  | Patients (N=45) | Percentage (%) |
| --- | --- | --- |
| **Sex** |  |  |
| Male | 19 | 42.2 |
| Female | 26 | 57.8 |
| **Age** | 64.4 ± 11.7 |  |
| **Underlying disease** |  |  |
| Diabetes mellitus | 13 | 28.9 |
| Hypertension | 21 | 46.7 |
| Cerebrovascular accident | 6 | 13.3 |
| Chronic kidney disease | 2 | 4.4 |
| Chronic obstructive pulmonary disease | 1 | 2.2 |
| **Type of cancer** |  |  |
| Intra-abdominal | 14 | 31.1 |
| Genitourinary | 24 | 53.3 |
| Lung | 4 | 8.9 |
| Others | 3 | 6.7 |
| **APACHE III** | 93.9 ± 29.2 |  |
| **SOFA** | 13.2 ± 3.3 |  |
| **No. of PMX-DHP\* treatment** |  |  |
| #1 | 38 | 84.4 |
| > #2 | 7 | 15.6 |
| **VIS** | 289.7 ± 220.5 |  |
| **Primary infection site** |  |  |
| Lung | 9 | 20 |
| Abdominal | 29 | 64.4 |
| Genitourinary | 5 | 11.1 |
| Others | 2 | 4.4 |

**Laboratory values**
|  |  |
| --- | --- |
| ANC (/uL) | 1575.5 (0–30589) |
| Platelet count (10<sup>3</sup>/uL) | 107 (2–299) |
| CRP<sup>#</sup> (mg/dL) | 15.5 (0.1–30.5) |
| Lactic acid (mg/dL) | 95 (13.6–353.4) |
| Procalcitonin (mg/mL) | 30.2 (0.4–526.6) |
| Creatinine (mg/dL) | 1.5 (0.3–7.7) |
| P/F ratio | 141.2 (20.2–534.3) |
| **Positive culture** |  |  |
| G(-) | 25 | 62.5 |
| G(+) | 11 | 27.5 |
| Mixed | 4 | 10 |
| **Mechanical ventilation** | 41 | 91.1 |
| **Intervention** | 14 | 68.9 |
| **Operation** | 16 | 64.4 |
| **28-day mortality** |  |  |
| Alive | 21 | 46.7 |
| Death | 24 | 53.3 |

ANC: absolute neutrophil count; APACHE III: Acute Physiology and Chronic Health Evaluation III (APACHE III); CRP: C-reactive protein; G(-): Gram-negative bacteremia; G(+): Gram-positive bacteremia; P/F ratio: ratio of arterial oxygen partial pressure to fraction of inspired oxygen (PaO<sub>2</sub>/FiO<sub>2</sub>); PMX-DHP: polymyxin B-immobilized fiber column direct hemoperfusion; SOFA: sequential organ failure assessment; VIS: vasoactive inotrope score (dopamine dose (μg/kg/min) + dobutamine dose (μg/kg/min) + 100 × epinephrine dose
( $\mu\text{g/kg/min}$ ) + $10 \times$ milrinone dose ( $\mu\text{g/kg/min}$ ) + $10,000 \times$ vasopressin dose (units/kg/min) + $100 \times$ norepinephrine dose ( $\mu\text{g/kg/min}$ )

When patients were divided into two groups— early and late —based on the PMX-DHP initiation time, the most frequent cancer type in both groups was genitourinary cancer (early *vs.* late, 55.0% *vs.* 52.0%, respectively). The disease severity indices (i.e., median APACHE III and SOFA scores) did not vary significantly between the two groups (APACHE III score: 95.5 (41–144) *vs.* 94 (36–159), p=0.427; SOFA score: 13 (6–18) *vs.* 14 (7–21), *p*=0.270). The VIS was higher in the early group than the late group (372.5 [15–913] *vs.* 213 [9–847], *p*=0.020). In both groups, the abdominal area was the most frequent primary infection site (60% *vs.* 68%). Gram-negative bacteremia was the most commonly detected bacteria on microbiological culture in both groups (64.7% *vs.* 60.9%, p=0.309). Compared to that of the early initiation group, the late initiation group showed a slightly higher rate of gram-positive bacteremia (17.6% *vs.* 34.8%), a higher rate of intervention to infection source control (15% *vs.* 44%, *p*=0.037), a lower frequency of surgical intervention (45% *vs.* 25%, *p*=0.237), and a higher level of 28-day mortality (30% *vs.* 60%, *p*=0.045) (Table 2).

**Table 2.** Baseline characteristics of the early *vs.* the late initiation treatment group.

|  | Early (N=20) | Late (N = 25) | *p*-value |
| --- | --- | --- | --- |
| **Sex** |  |  |  |
| Male | 9 (45.0%) | 10 (40.0%) | 0.736 |
| Female | 11 (55.0%) | 15 (60.0%) |  |
| **Age** | 66.2 ± 11.2 | 62.9 ± 12.0 | 0.362 |
| **Underlying disease** |  |  |  |
| Diabetes mellitus | 13 (65.0%) | 19 (76.0%) | 0.419 |
| Hypertension | 11 (55.0%) | 10 (40.0%) | 0.316 |
| Cerebrovascular accident | 5 (25.0%) | 1 (4.0%) | 0.074 |
| Chronic kidney disease | 1 (5.0%) | 1 (4.0%) | 1 |
| Chronic obstructive pulmonary disease | 1 (5.0%) | 0 (0.0%) | 0.444 |
| **Type of cancer** |  |  |  |
| Intra-abdominal | 5 (25.0%) | 9 (36.0%) | 0.792 |
| Genitourinary | 11 (55.0%) | 15 (52.0%) |  |
| Lung | 2 (10.0%) | 2 (8.0%) |  |
| Others | 2 (10.0%) | 1 (4.0%) |  |
| **APACHE III** | 97.8 ± 30.8 | 90.7 ± 28.1 | 0.427 |
| **SOFA** | 12.6 ± 3.2 | 13.7 ± 3.4 | 0.27 |
| **No. of PMX-DHP treatment** |  |  |  |
| #1 | 17 (85.0%) | 21 (84.0%) | 1 |
| > #2 | 3 (15.0%) | 4 (16.0%) |  |
| **VIS** | 365.9 ± 207.2 | 228.8 ± 215.5 | 0.02 |
| **Primary infection site** |  |  |  |
| Lung | 3 (15.0%) | 6 (24.0%) | 0.411 |
| Abdominal | 12 (60.0%) | 17 (68.0%) |  |
| Genitourinary | 4 (20.0%) | 1 (4.0%) |  |
| Others | 1 (5.0%) | 1 (4.0%) |  |
| **Positive culture** |  |  |  |
| G(-) | 11 (64.7%) | 14 (60.9%) | 0.309 |
| G(+) | 3 (17.6%) | 8 (34.8%) |  |
| Mixed | 3 (17.6%) | 1 (4.3%) |  |
| **Mechanical ventilation** | 19 (95.0%) | 22 (88.0%) | 0.617 |
| **Intervention** | 3 (15.0%) | 11 (44.0%) | 0.037 |
| **Operation** | 9 (45.0%) | 7 (28.0%) | 0.237 |
| **28-day mortality** |  |  |  |
| Alive | 14 (70.0%) | 10 (40.0%) | 0.045 |
| Death | 6 (30.0%) | 15 (60.0%) |  |
| **ANC (uL)** | 1191 (0–22568) | 2266 (0–30589) | 0.097 |
| Non-Neutropenia (ANC>1500) | 9 (45%) | 15 (62.5%) | 0.246 |
| Neutropenia (ANC<1500) | 11 (55.0%) | 9 (37.5%) |  |
| **Platelet count (10<sup>3</sup>/uL)** | 117.5 (13–299) | 96 (2–246) | 0.289 |
| **CRP (mg/dL)** | 14.9 (0.1–27.3) | 15.8 (1.1–30.5) | 0.173 |
| **Lactic acid (mg/dL)** | 90.3 (13.6–353.4) | 95 (26.1–262.9) | 0.807 |
| **Procalcitonin (mg/mL)** | 44.5 (0.4–144.2) | 20.7 (1.3–526.6) | 0.34 |
| **Creatinine (mg/dL)** | 1.4 (0.3–7.7) | 1.5 (0.4–4) | 1 |
| **P/F ratio** | 121.8 (20.2–524) | 156 (48.1–534.3) | 0.576 |

ANC: absolute neutrophil count; APACHE III: Acute Physiology and Chronic Health Evaluation III (APACHE III); CRP: C-reactive protein; G (-): Gram-negative bacteremia; G (+): Gram-positive bacteremia; P/F ratio: ratio of arterial oxygen partial pressure to fraction of inspired oxygen (PaO<sub>2</sub>/FiO<sub>2</sub>); PMX-DHP: polymyxin B-immobilized fiber column direct hemoperfusion; SOFA: sequential organ failure assessment (SOFA); VIS: vasoactive inotrope score

### Overall survival in the early vs. late group

The number of deaths within 28 days from the initiation of PMX-DHP was compared for both groups; six deaths were recorded in the early and 15 in the late initiation group (Table 2). The survival rate was ≥50% in the early group, and the median survival was 7 days in the late group. While the 28-day overall survival was slightly higher in the early compared to the late group, no significant difference was found (*p*=0.0515) (Fig 1).

**Fig 1.**
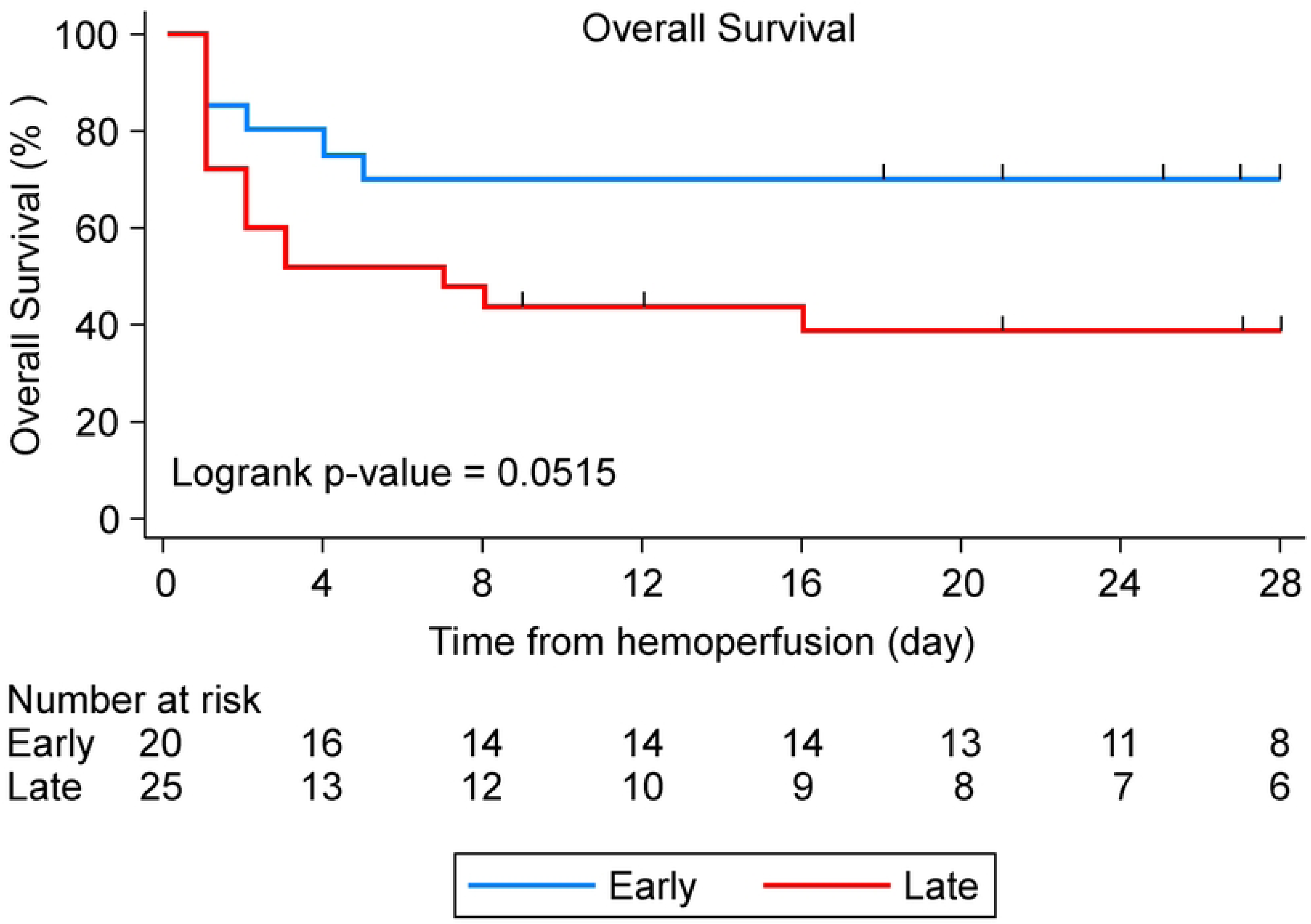
Estimation of overall survival according to initiation of polymyxin B-immobilized fiber column direct hemoperfusion (PMX-DHP).

### Univariable and multivariable analyses of 28-day mortality

Our univariable analysis of 28-day mortality in patients with cancer with refractory septic shock after PMX-DHP showed that DM (odds ratio (OR)=3.081; 95% confidence interval (CI)=1.290–7.360; *p*=0.011), lactic acid levels (OR=1.010; 95% CI=1.005–1.014; *p*<0.0001), and SOFA scores (OR=1.190; 95% CI=1.044–1.357; *p*=0.009) were correlated with 28-day mortality. A multivariable analysis was performed using backwards selection and the univariable analysis results; the results indicated that 28-day mortality was correlated with DM (OR=2.718; 95% CI=1.013–7.291; *p*=0.047), early initiation (OR=0.268; 95% CI=0.094–0.765; *p*=0.014), and lactic acid levels (OR=1.009; 95% CI=1.004–1.014; *p*<0.0001) (Table 3).

**Table 3.** Univariable and multivariable analysis of 28-day mortality.

|  | Univariable analysis |  | Multivariable analysis |  |
| --- | --- | --- | --- | --- |
|  | OR (95% CI) | *p*-value | OR (95% CI) | *p*-value |
| Sex | 1.294 (0.549–3.049) | 0.555 |  |  |
| **Age** | 1.001 (0.065–1.038) | 0.974 |  |  |
| **Underlying disease** |  |  |  |  |
| Diabetes mellitus | 3.081 (1.290–7.360) | 0.011 | 2.718 (1.013–7.291) | 0.047 |
| Hypertension | 1.098 (0.466–2.587) | 0.83 |  |  |
| Cerebrovascular<br>accident | 1.206 (0.3555–4.097) | 0.765 |  |  |
| Chronic kidney<br>disease | 4.025 (0.898–18.042) | 0.069 |  |  |
| COPD | 3.126 (0.409–23.917) | 0.272 |  |  |
| **Initiation of PMX-DHP** |  |  |  |  |
| Late |  |  |  |  |
| Early | 0.423 (0.163–1.093) | 0.076 | 0.268 (0.094–0.765) | 0.014 |
| **Type of cancer** |  |  |  |  |
| Intra-abdominal | 2.171 (0.274–17.166) | 0.463 |  |  |
| Genitourinary | 0.948 (0.119–7.578) | 0.96 |  |  |
| Lung | 2.205 (0.229–21.199) | 0.494 |  |  |
| Others |  |  |  |  |
| **APACHE III** | 1.002 (0.988–1.017) | 0.77 |  |  |
| **SOFA score** | 1.190 (1.044–1.357) | 0.009 |  |  |
| **VIS** | 1.002 (1.000–1.004) | 0.093 |  |  |
| **Primary infection site** |  |  |  |  |
| Lung | 1.258 (0.151–10.46) | 0.832 |  |  |
| Abdominal | 0.696 (0.090–5.362) | 0.728 |  |  |
| Genitourinary | 0.697 (0.063–7.694) | 0.768 |  |  |
| **Mechanical ventilation** | 5.373 (0.303–95.258) | 0.252 |  |  |
| **Intervention** | 1.138 (0.459–2.821) | 0.78 |  |  |
| **Operation** | 0.474 (0.173–1.298) | 0.146 |  |  |
| **ANC (/uL)** |  |  |  |  |
| Non-Neutropenia (ANC>1500) |  |  |  |  |
| Neutropenia (ANC<1500) | 0.560 (0.223–1.408) | 0.218 |  |  |
| **Platelet count (10<sup>3</sup>/uL)** | 0.996 (0.990–1.002) | 0.194 |  |  |
| **CRP (mg/dL)** | 0.992 (0.938–1.049) | 0.769 |  |  |
| **Lactic acid (mg/dL)** | 1.010 (1.005–1.014) | <0.0001 | 1.009 (1.004–1.014) | <0.0001 |
| **Procalcitonin (mg/mL)** | 0.990 (0.973–1.008) | 0.281 |  |  |
| **Creatinine (mL/dL)** | 1.165 (0.911–1.490) | 0.223 |  |  |
| **P/F ratio** | 0.998 (0.005–1.002) | 0.343 |  |  |

ANC: absolute neutrophil count; APACHE III: Acute Physiology and Chronic Health Evaluation III (APACHE III); COPD: chronic obstructive pulmonary disease; CRP: C- reactive protein; P/F ratio: ratio of arterial oxygen partial pressure to fraction of inspired oxygen (PaO<sub>2</sub>/FiO<sub>2</sub>); PMX-DHP: polymyxin B-immobilized fiber column direct hemoperfusion; SOFA: sequential organ failure assessment (SOFA); VIS: vasoactive inotrope score; OR: odds ratio; CI: confidence interval.

### Changes in values of variables after PMX-DHP treatment (early vs. late initiation)

Comparison of variables before and 24 h after PMX-DHP showed that, in both groups, VIS (p=0.001 *vs.* p=0.005, respectively) and creatinine levels (*p*=0.016 *vs. p*=<0.001) significantly decreased after PMX-DHP. However, the decrease in the VIS was higher in the early initiation group (Fig 2).

**Fig 2.**
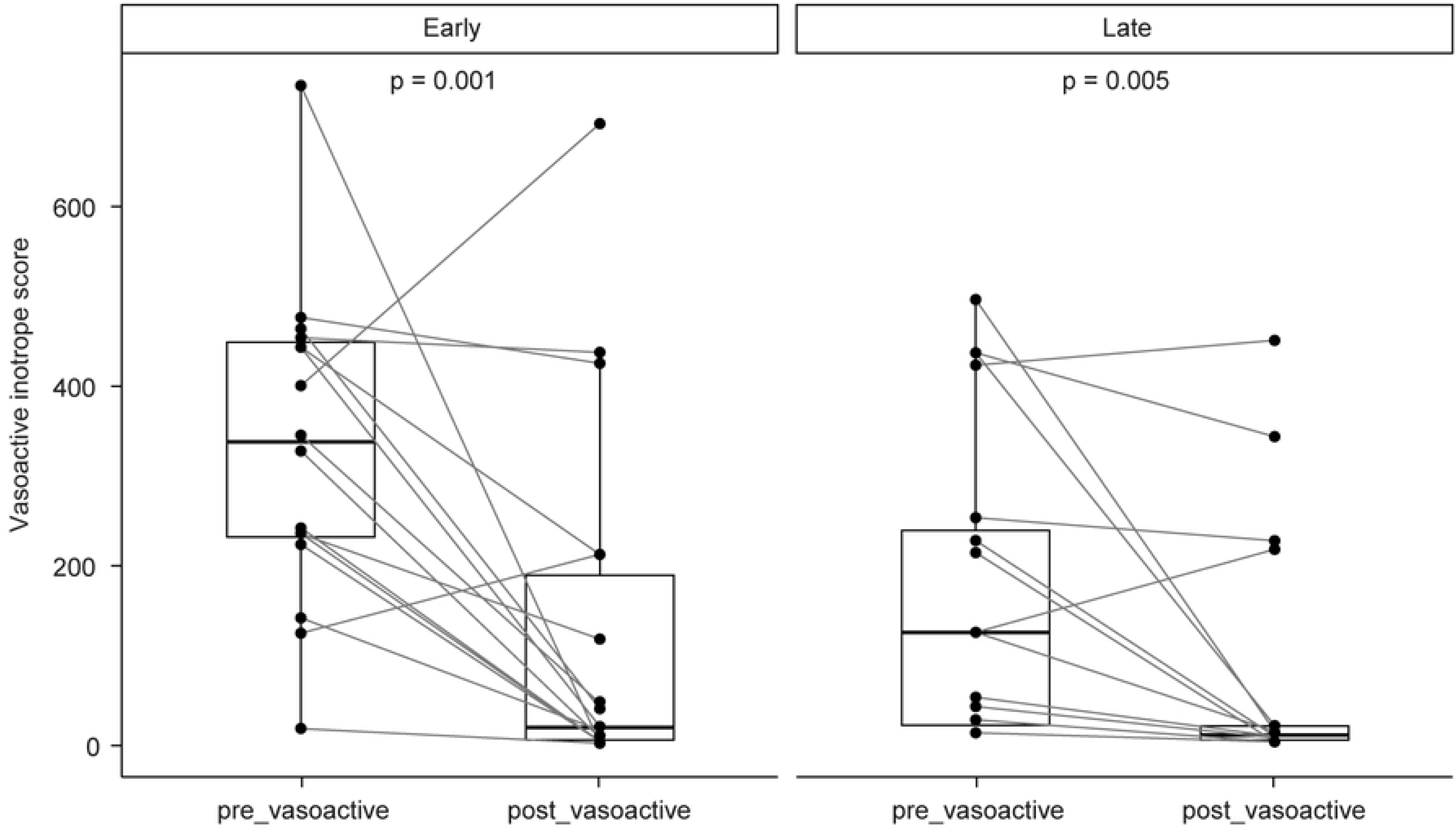
Changes in variables associated with initiation of polymyxin B-immobilized fiber column direct hemoperfusion (PMX-DHP): vasoactive inotrope score (VIS)

In both groups, the platelet count (Δ62.0 *vs.* Δ25.5; *p*=0.007 *vs. p*=0.061, respectively), lactic acid levels (Δ37.5 *vs.* Δ33.2; *p*=0.080 *vs. p*=0.083), and SOFA scores (*p*=0.230 *vs. p*=0.022) significantly decreased after PMX-DHP. After 24 h, the PaO_2_/FiO_2_ ratio was higher in both groups (Δ127.5 *vs.* Δ95.6; *p*=0.350 *vs. p*=0.390, respectively), with the degree of increase being slightly higher in the early initiation group (Table 4).

**Table 4.** Changes in variables after PMX-DHP treatment.

|  | Before HP | After HP | *p*-value |
| --- | --- | --- | --- |
| **Early initiation** |  |  |  |
| ANC (/uL) | 1191 (0–22568) | 3158.5 (0–23000) | 0.083 |
| Platelet count ($10^3/\text{uL}$) | 117 (13–299) | 55 (16–165) | 0.007 |
| CRP (mg/dL) | 14.5 (0.1–27.3) | 18.5 (8.2–38) | 0.015 |
| Lactic acid (mg/dL) | 73.7 (13.6–353.4) | 36.2 (17.3–278.5) | 0.080 |
| Procalcitonin (mg/mL) | 44.5 (0.4–144.2) | 54.7 (19.1–307.3) | 0.400 |
| Creatinine (mg/dL) | 1.3 (0.3–6.6) | 1.1 (0.3–2.6) | 0.016 |
| P/F ratio | 160 (20.2–524) | 287.5 (37.2–498) | 0.350 |
| SOFA score | 11 (6–17) | 10 (4–15) | 0.230 |
| VIS | 336 (15–736) | 18 (0–693) | 0.001 |
| **Late initiation** |  |  |  |
| ANC (/uL) | 2214.5 (1–30589) | 11110.5 (4–35040) | 0.830 |
| Platelet count (10<sup>3</sup>/uL) | 116 (3–246) | 90.5 (0–138) | 0.061 |
| CRP (mg/dL) | 17.6 (4.4–30.5) | 18.4 (2.3–39) | 0.920 |
| Lactic acid (mg/dL) | 82.4 (26.1–234.8) | 49.2 (13–287.5) | 0.083 |
| Procalcitonin (mg/mL) | 22.7 (1.3–526.6) | 19.7 (0.1–238.7) | 0.110 |
| Creatinine (mg/dL) | 1.5 (0.5–3.7) | 1 (0.5–1.8) | <0.0001 |
| P/F ratio | 164.9 (48.1–134.3) | 260.5 (66.6–544.4) | 0.390 |
| SOFA score | 13 (7–21) | 12 (3–18) | 0.022 |
| VIS | 124 (9–496) | 10 (0–451) | 0.005 |

ANC: absolute neutrophil count; CRP: C-reactive protein; P/F ratio: ratio of arterial oxygen partial pressure to fraction of inspired oxygen (PaO<sub>2</sub>/FiO<sub>2</sub>); PMX-DHP: polymyxin B-immobilized fiber column direct hemoperfusion; SOFA: sequential organ failure assessment (SOFA); VIS: vasoactive inotrope score

## Discussion

This study was conducted to analyze the correlation between PMX-DHP initiation time and 28-day mortality in patients with cancer with refractory septic shock and to verify whether hemodynamics, SOFA scores, and oxygenation levels improve after PMX-DHP. A mortality rate of 53.3% related to refractory septic shock was observed in this study, which did not significantly differ from that of a previous meta-analysis of patients with cancer after sepsis treatment (48–62%) [13]. However, the earlier reported mortality has been found to be slightly higher than the 28-day mortality related to septic shock observed in our patients with cancer who received PMX-DHP (19–37.7%) [7,8,14]. This may be due the immunosuppressed nature of our patients which resulted in higher disease severity (APACHE III score; SOFA score) than those in studies performed at other centers. Another possible explanation could be that the treatment initiation in the present study occurred at a comparatively more severe stage, as the initiation of PMX-DHP follows the diagnosis of refractory septic shock.

The results of this study demonstrate that the requirement for a vasoactive inotrope agent to maintain hemodynamics after PMX-DHP decreased in the early initiation group, which led to a fall in 28-day mortality. The inotropes-vasopressor requirement decreased (VIS, Δ318 *vs.* Δ114, *p*=0.001 *vs. p*=0.005) after PMX-DHP in both the early and the late group; however, the decrease was larger in the early than the late group. Before the treatment, the requirement for a vasoactive inotrope agent to maintain hemodynamics was higher in the early than the late group; this enabled a relatively faster detection of refractory septic shock by the attending physician and thereby the administration of PMX-DHP, which reduced 28-day mortality. This implies that rapid administration of PMX-DHP within 12 h could improve hemodynamics and prevent life-threatening organ failure, thereby contributing to a reduction in mortality.

Through the modulation of activated mononuclear cells and neutrophils, PMX-DHP improves pulmonary oxygenation by suppressing inflammatory mediators and systemic inflammation [15]. In the current study, the PaO_2_/FiO_2_ ratio was higher in both groups after PMX-DHP, with a slightly larger increase in the early initiation group, although the difference was not statistically significant (Δ127.5 *vs.* Δ95.6; *p*=0.350 *vs. p*=0.390). The PaO_2_/FiO_2_ ratio was assessed 24 h after PMX-DHP in the current study; this fact might explain the differences between ours and the findings of previous studies, where the target was acute respiratory distress syndrome or acute lung injury and the PaO2/FiO2 ratio was evaluated 72–96 h after PMX-DHP [5,10,15].

Studies have shown that PMX-DHP prevents acute kidney injury and renal tubular cell apoptosis by maintaining the acid-base balance and lowering the plasma concentrations of lactate, creatinine, IL-6, and IL-10 [16]. In the current study, likewise, the creatinine level decreased in both groups after PMX-DHP (early group, *p*=0.016 *vs.* late group, *p*<0.001). However, this could not be confirmed to be an outcome of PMX-DHP, as most patients (78%) also underwent continuous renal replacement therapy. In addition, studies where continuous renal replacement therapy and PMX-DHP were concurrently applied did not report a reduction in mortality [17]. Moreover, creatinine levels did not significantly affect 28-day mortality in our patient sample, which corresponds with the results of previous studies.

Conventionally, the duration of PMX-DHP is 2 h; however, recent studies have reported that, in the event of endotoxin adsorption saturation, extending its duration to >2 h could enhance pulmonary oxygenation and hemodynamics, thereby reducing 28-day mortality [18–20]. The PMX-DHP duration in the current study was 15 h on an average. One or two additional treatments were performed by the attending physician; however, no significant correlation was found between the treatment frequency and 28-day mortality (OR=1.640; 95%CI=0.598–4.499; *p*=0.036).

The most common adverse effects of PMX-DHP are thrombocytopenia, transient hypotension, and allergic reactions [21]. Accordingly, the platelet count after hemoperfusion in the current study decreased in both the early and late initiation groups (Δ62 *vs.* Δ25.5; *p*=0.007 *vs. p*=0.061). The patients in our sample did not have transient hypotension or allergic reactions.

This study has certain limitations. First, this was a single-center, retrospective study with a small sample size, which limits the generalizability of our findings. Second, we focused on patients with refractory septic shock without initial measurements of endotoxin levels, which limited our analysis of endotoxemia-targeted PMX-DHP treatment effects. However, PMX-DHP is known to induce the absorption of endogenous cannabinoids, activated neutrophils, and monocytes, modify the monocyte surface marker expression, and be an effective treatment option for gram-positive bacteremia [15,22]. We can therefore infer that PMX-DHP provides beneficial effects for both gram-negative and positive bacteremia.

Despite these limitations, this is the first study to assess the effects of PMX-DHP in patients with cancer with refractory septic shock, and our findings confirm that the early initiation of PMX-DHP, compared to its late initiation, can reduce 28-day mortality and the inotropes-vasopressor requirement for maintaining hemodynamics. In the future, a multicenter, large-scale, randomized, controlled study should be conducted to determine the optimal initiation time.

## Conclusions

In our sample of patients with cancer with refractory septic shock, the early initiation of PMX-DHP, compared to its late initiation, was effective in reducing 28-day mortality and the inotropes-vasopressor requirement for maintaining hemodynamics. Based on these findings, the optimal initiation time should be determined in a follow-up study.

## Acknowledgements

The author thanks Dr. Jee hee-Kim for invaluable help and advice throughout the study and Ms. Mira Han for assistance with bioinformatics and statistics; and Editage for English language editing.

## Data availability

All data generated or analyzed during this study are included in this published article.

## Notes

### Competing Interest Statement

The authors have declared that no competing interests exist.

### Funding Statement

This work was supported by the National Cancer Center, Korea (NCC grant: 2212490-2). The funders had no role in study design, data collection and analysis, decision to publish, or preparation of the manuscript.

### Author Declarations

This study was approved by the Institutional Reviewer Board of the National Cancer Center(Approval No,: NCC 2023-0170).

